# Prediction of Uterine Dehiscence via Machine Learning by Using Lower Uterine Segment Thickness and Clinical Features

**DOI:** 10.1101/2022.03.23.22272815

**Authors:** Mervenur Kement, Cihan Emre Kement, Mahmut Kuntay Kokanali, Melike Doganay

## Abstract

**Background:** With the global increase of Cesarean section delivery rates, the long-term effects of Cesarean delivery have started to become more clear. One of the most prominent complications of Cesarean section in recurrent pregnancies is uterine rupture. Assessing the risk of uterine rupture or dehiscence is very important in order to prevent untimely operations and/or maternal and fetal complications.

**Objective:** Our study aims to assess whether machine learning can be used to predict uterine dehiscence or rupture by using patients’ ultrasonographic findings, clinical findings and demographic data as features. Hence, possible uterine rupture, as well as maternal and fetal complications pertinent to it, could be prevented.

**Study Design:** The study was conducted on 317 patients with term (>37 weeks) singleton pregnancy. Demographics, body-mass indices, smoking and drinking habits, clinical features, past pregnancies, number and history of abortions, inter-delivery period, gestation week, number of previous Cesarean operations, fetal presentation, fetal weight, tocography data, trans-abdominal ultrasonographic measurement of lower uterine segment full thickness and myometrium thickness, lower uterine segment findings during Cesarean section were collected and analyzed using machine learning techniques. Logistic Regression, Multilayer Perceptron, Support Vector Machine, Random Forest and Naive Bayes algorithms were used for classification. The dataset was evaluated using 10-fold cross-validation. Correct Classification Rate, F-score, Matthews Correlation Coefficient, Precision-Recall Curve area and Receiver Operating Characteristics area were used as performance metrics.

**Results:** Among the machine learning techniques that has been tested in this study, Naive Bayes algorithm showed the best prediction performance. Among the various combinations of features used for prediction, the essential features of parity, gravida, tocographic contraction, dilation, d&c with the sonographic thickness of lower uterine segment myometrium yielded the best results. The runner-up performance was obtained with the sonographic full thickness of lower uterine segment added to the base features. The base features alone can classify patients with 90.5% accuracy, while adding the myometrium measurement increases the classification performance by 5.1% to 95.6%. Adding the full thickness measurement to the base features raises the classification performance by 4.8% to 95.3% in terms of Correct Classification Rate.

**Conclusion:** Naive Bayes algorithm can correctly classify uterine rupture or dehiscence with a Correct Classification Rate of 0.953, an F-score of 0.952 and a Matthews Correlation Coefficient value of 0.641. This result can be interpreted such that by using clinical features and lower uterine segment ultrasonography findings, machine learning can be used to accurately predict uterine rupture or dehiscence.

**Trial registration:** Clinical Research Ethics Committee of Ankara City Hospital, University of Health Sciences (Approval number: E2-20-108)

Date of registration: 27-01-2021

URL: https://ankarasehir.saglik.gov.tr/TR-348810/2-nolu-etik-kurul.html

**AJOG at a Glance:** *Why was this study conducted?:* This study was conducted to: - Determine whether machine learning algorithms can be utilized to predict uterine dehiscence and assess the risk of uterine rupture
- Evaluate the contribution of ultrasonographic measurement of lower uterine segment measurements to the prediction performance of the algorithms
- Find out which machine learning technique performs the best for predicting uterine dehiscence.

*What are the key findings?:*

- Machine learning methods can be used to accurately predict uterine dehiscence (with up to 95.6% accuracy).
- Using lower uterine segment full thickness or myometrium thickness increases the accuracy of Naive Bayes algorithm by 4.8% and 5.1%, respectively.
- Naive Bayes algorithm yields the best prediction performance among the methods tried.

*What does this study add to what is already known?:*

- Ultrasonographic lower uterine segment measurements can be used as features in machine learning to increase its prediction performance of uterine dehiscence and hence the risk of uterine rupture.

## 1. Introduction

The concern for uterine rupture can cause the patients with contractions and with previous Cesarean delivery to go under preterm Cesarean operation. Repeat Cesarean not only increases the risk of uterine rupture, but also neonatal complications such as bleeding, hysterectomy, thromboembolism and respiratory distress syndrome [1, 2]. The risk is increased for preterm babies [3]. Therefore, being able to accurately predict uterine dehiscence is of great importance in order to prevent untimely Cesarean operations.

When a preterm Cesarean decision is made, presence of contraction in tocography is used as an important indicator by the gynecologist. However, a significant portion of such patients are observed to have no uterine dehiscence or rupture during the operation. One study [4] used the lower uterine segment (LUS) ultrasound measurements to predict uterine dehiscence or rupture. An earlier study by Rozenberg et al. [5] showed that uterine defects are directly correlated with the thinning of LUSs. The study deduces that the prevalence of uterine scar defects is 16% in patients with LUS thickness of less than 2.5mm, whereas it is 0.7% for patients with LUS thickness of more than 3.5mm. Other studies also correlate thin LUS with increased risk of rupture [6–14]. On the other hand, meta-analyses indicate that LUS alone is not sufficient to predict uterine rupture, while concurring that LUS thickness less than 2 millimeters pose a risk for rupture and dehiscence [15, 16].

In this study we aim to assess whether uterine dehiscence or rupture can be predicted via machine learning methods by using LUS measurements as well as clinical findings and demographics of patients as features. Moreover, we aim to find out which of these features are more relevant to the prediction that machine learning algorithms try to make. Furthermore, we compare the effects of using LUS myometrium thickness and LUS full thickness as features, as well as the impact of using these two features together on the prediction performance of machine learning (ML) techniques.

## 2. Materials and Methods

### Study Design and Participants

The study was approved by Clinical Research Ethics Committee of Ankara City Hospital, University of Health Sciences (Approval number: E2-20-108) in January 2021. Singleton pregnancies who have had cesarean operation and applied to the Department of Obstetrics and Gynecology Labor Unit of Ankara City Hospital, Ankara, Turkey between February and July 2021 were included in this study. The study is conducted on 317 patients with term (>37 weeks) singleton pregnancy. Information regarding demographics, body-mass indices, smoking and drinking habits, clinical features, past pregnancies, number and history of abortions, inter-delivery period, gestation week, number of previous cesarean operations, fetal presentation, fetal weight, tocography data, trans-abdominal ultrasonographic measurement of lower uterine segment full thickness and myometrium thickness, lower uterine segment findings during cesarean section was collected[17]. 24 patients were excluded from the study since LUS could not be screened properly due to bladder adhesions. Transabdominal sonographic examination was carried out with a full urinary bladder to allow good imaging of the LUS. All examinations were performed on an Voluson GE S10 series 2-4 MHz probe by a single sonographer. The patients’ labor and delivery outcomes were reviewed and any evidence of rupture or dehiscence were noted. 9 additional patients were excluded from the study due to previous incisions that were not transverse.

### Statistical Analysis

Statistical analysis was performed with Mann-Whitney U, chi-square and Fisher exact tests. *P* < 0.05 was taken as significant. In order to increase the performance of ML methods, in the data preprocessing stage we eliminated the features that were found to be less relevant to uterine defects. While determining the features to be omitted, we used the statistical significance as well as the Information Gain and Gain Ratio metrics [18]. Out of the 22 features collected, we selected 10 features based on their statistical significance, Information Gain ranking and Gain Ratio ranking. Since different ML algorithms perform better than the others in different applications; Logistic Regression, Multilayer Perceptron, Support Vector Machine, Random Forest and Naive Bayes algorithms [19] were all trained and tested in this study to assess which alhorithm outperforms the others. Since the dataset is relatively small and the prevalence of uterine dehiscence and rupture in general is relatively low, the data was not divided into training and validation sets. Instead, the performance of the algorithms was measured by using 10-fold cross-validation. Multiple performance criteria were selected as Correct Classification Rate (CCR), F-score, Matthews Correlation Coefficient (MCC), Precision-Recall Curve (PRC) area and Receiver Operating Characteristics (ROC) area in order to accurately measure and compare the performances.

## 3. Results

The mean age and mean BMI of the patients were found to be 28.74±4.93 and 29.22±4.41, respectively. 11.1% of the patients reported to be smokers. The number of patients with gravida ≤3 was 245 (77.3%). 277 of the patients had parity ≤2. The mean number of abortions were 0.30 ± 0.67 whereas 14 (4.4%) of the patients had dilation & curettage. Gestation week was ≤ 39 for the 77.9% of the patients and > 39 for the 22.1% of the patients.

183 (57.7%) patients had contractions in Cardiotocography (CTG) and the mean value in Montevideo units (MVU) was 121.72 ± 33.59. Full LUS measurements were < 1*mm* for 4 (1.3%), 1−2*mm* for 45 (14.2%), and ≥ 2*mm* for 268 (84.5%) of the patients. Estimated fetal weight was ≤ 3000*gr* for 90 (28.4%) of the patients and > 3000*gr* for 227 (71.6%) of the patients. Lastly, intraoperative dehiscence was observed in 23 (7.3%) of the patients.

Table 2 shows the quantitative variables’ means, standard deviations and median values in case of dehiscence and no dehiscence. Statistical significance was observed for age, number of previous Cesareans, number of years passed since first Cesarean and effacement features (p values are < 0.001, < 0.001, < 0.001 and 0.034, respectively). The mean age and the mean number of previous Cesareans of the patients with uterine dehiscence is significantly larger than the patients with no uterine dehiscence. Furthermore, patients with no dehiscence had their first Cesarean on average 5.80 ± 3.59 years before, while patients with dehiscence had their first Cesarean 9.78 ± 4.06 years before on average. Moreover, effacement was also significantly larger in patients with dehiscence (24.35±21.07) than in patients with no dehiscence (15.37 ± 20.33).

Qualitative variables regarding the patients were presented in Table 1 in case of intraoperative dehiscence and no intraoperative dehiscence. Gravida, parity, contractions in CTG, dilation, LUS full measurement and LUS myometrium measurement were found to be statistically significant (p values are < 0.001, < 0.001, 0.038, 0.034, < 0.001 and < 0.001, respectively). Among the group of patients with dehiscence, 69.6% was found to have a gravida more than 3, and 60.9% was found to have a parity more than 2. Moreover, 78.3% demonstrated contraction in CTG, 60.9% had dilation, 82.6% had LUS full measurement between 1-2mm, and 95.7% had LUS full measurement less than 1mm.

**Table 1:** Quantitative patient characteristics.

| Features | Intraoperative dehiscence |  |  |  | p |
| --- | --- | --- | --- | --- | --- |
|  | No |  | Yes |  |  |
|  | Mean±Std. deviation | Median | Mean±Std. deviation | Median |  |
| Age | 28.43±4.82 | 28.00 | 32.74±4.67 | 34.00 | <0.001 |
| BMI | 29.10±4.33 | 28.70 | 30.80±5.26 | 29.50 | 0.159 |
| Abortions | 0.28±0.66 | 0.00 | 0.48±0.79 | 0.00 | 0.179 |
| Abortions following Cesarean | 0.18±0.53 | 0.00 | 0.30±0.63 | 0.00 | 0.213 |
| D&Cs following Cesarean | 0.12±0.39 | 0.00 | 0.26±0.62 | 0.00 | 0.251 |
| Previous Cesareans | 1.34±0.56 | 1.00 | 2.48±0.99 | 3.00 | <0.001 |
| Years since first Cesarean | 5.80±3.59 | 5.00 | 9.78±4.06 | 9.00 | <0.001 |
| Contraction (per 10 min) | 120.99±33.72 | 120.00 | 128.33±32.58 | 120.00 | 0.346 |
| Monitor. time (min) | 110.38±53.09 | 120.00 | 114.78±59.76 | 120.00 | 0.831 |
| Effacement (%) | 15.37±20.33 | 0.00 | 24.35±21.07 | 30.00 | 0.034 |

**Table 2:** Qualitative patient characteristics.

| Features |  | Intraoperative dehiscence |  |  |  | p |
| --- | --- | --- | --- | --- | --- | --- |
|  |  | No |  | Yes |  |  |
|  |  | n | % | n | % |  |
| Smoking | No | 264 | 89.8 | 21 | 91.3 | 1.00 |
|  | Yes | 30 | 10.2 | 2 | 8.7 |  |
| Gravida | ≤3 | 238 | 81 | 7 | 30.4 | <0.001 |
|  | >3 | 56 | 19 | 16 | 69.6 |  |
| Parity | ≤2 | 268 | 91.2 | 9 | 39.1 | <0.001 |
|  | >2 | 26 | 8.8 | 14 | 60.9 |  |
| D&C | No | 283 | 96.3 | 20 | 87 | 0.072 |
|  | Yes | 11 | 3.7 | 3 | 13 |  |
| Gestation week | ≤39 | 228 | 77.6 | 19 | 82.6 | 0.573 |
|  | >39 | 66 | 22.4 | 4 | 17.4 |  |
| Years since previous CS | ≤2 | 69 | 23.5 | 3 | 13 | 0.250 |
|  | >2 | 225 | 76.5 | 20 | 87 |  |
| Contraction | No | 129 | 43.9 | 5 | 21.7 | 0.038 |
|  | Yes | 165 | 56.1 | 18 | 78.3 |  |
| Dilation | No | 181 | 61.6 | 9 | 39.1 | 0.034 |
|  | Yes | 113 | 38.4 | 15 | 60.9 |  |
| Presentation | Cephalic | 287 | 97.6 | 23 | 100 | 1.00 |
|  | Breech | 5 | 1.7 | 0 | 0 |  |
|  | Transverse | 2 | 0.7 | 0 | 0 |  |
| LUS full (mm) | ≤1 | 1 | 0.3 | 3 | 13 | <0.001 |
|  | 1-2 | 26 | 8.8 | 19 | 82.6 |  |
|  | >2 | 267 | 90.9 | 1 | 4.3 |  |
| LUS myometrium | ≤1 | 26 | 8.8 | 22 | 95.7 | <0.001 |
|  | 1-2 | 127 | 43.2 | 1 | 4.3 |  |
|  | >2 | 141 | 48 | 0 | 0 |  |
| Est. fetal weight (gr) | ≤3000 | 82 | 27.9 | 7 | 30.4 | 0.821 |
|  | >3000 | 212 | 72.1 | 16 | 69.6 |  |

Gain ratio and information gain results of the features were given in Figures 1 and 2, respectively. Note that all the features were not shown in these graphs, as the features with highest gain ratio and information gain scores were included in the graphics. Furthermore, note that the features in Figures 1 and 2 and the features that were found to be statistically significant do not overlap since p value, information gain and gain ratio are different metrics each of which gives an insight about the relationship between the features and uterine dehiscence.

**Figure 1:**
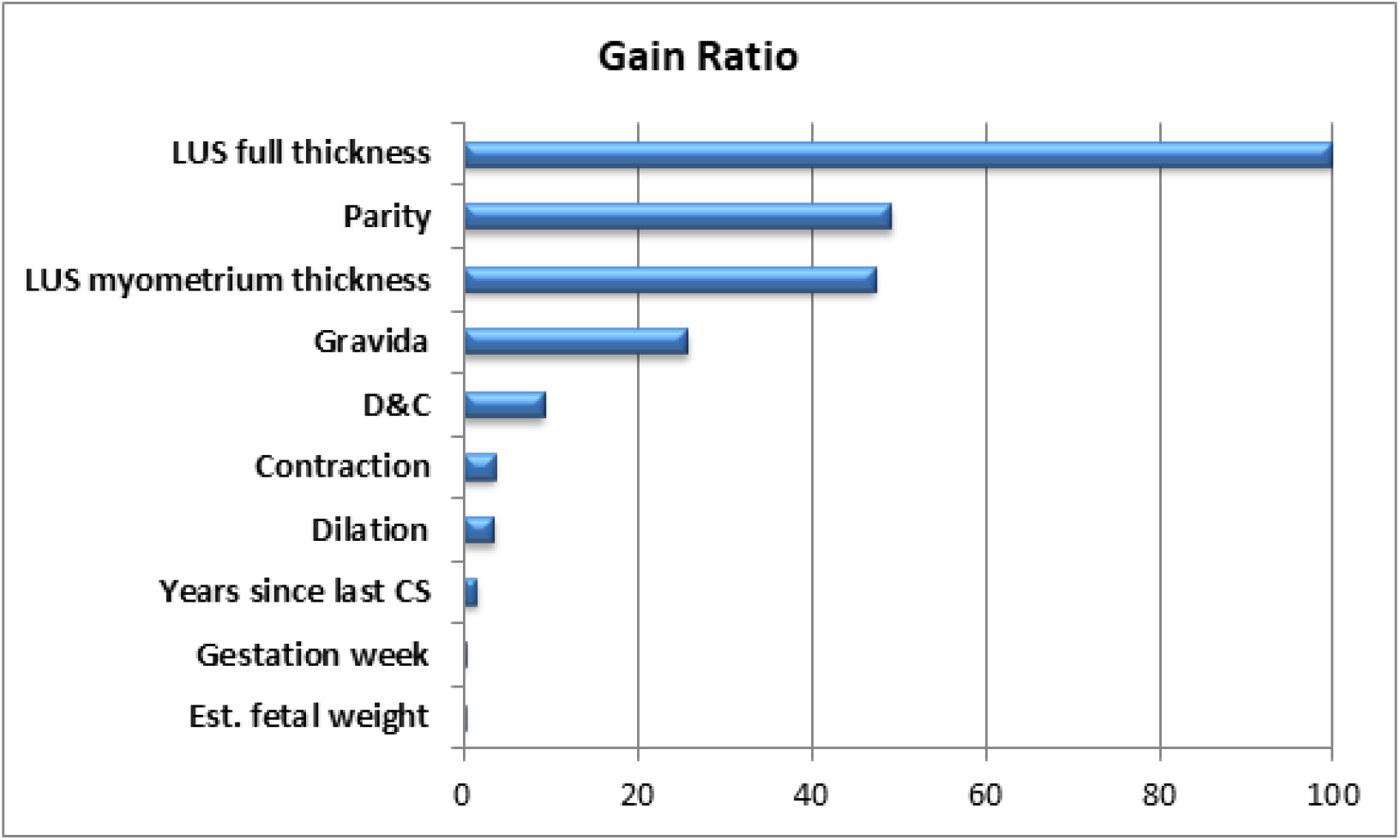
Gain ratio of features

**Figure 2:**
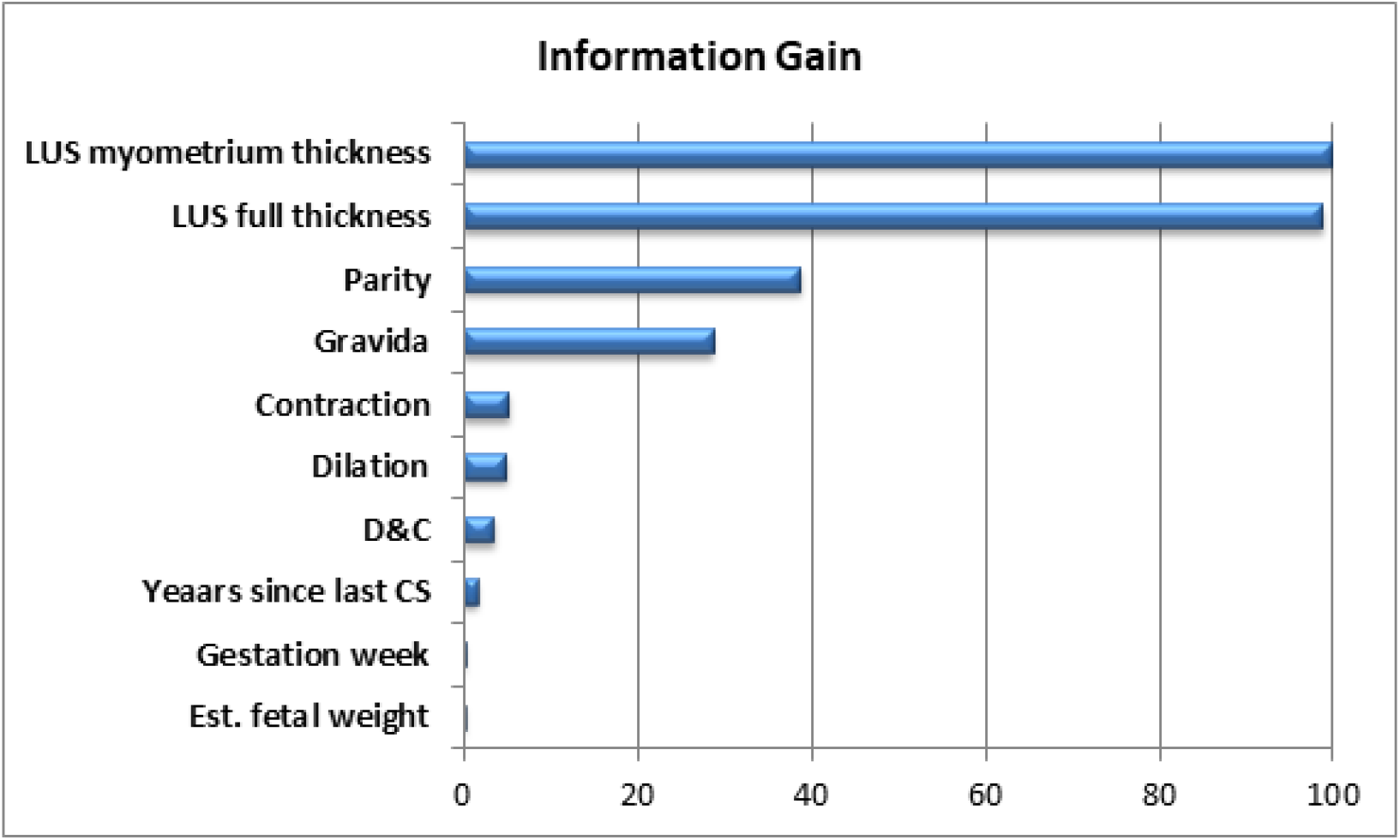
Information gain of features

The performance of the ML methods in terms of CCR (accuracy), F-score, MCC, PRC area and ROC area was given in Table 3. Note that the features included in this model are LUS myometrium thickness, parity, gravida, contraction, dilation, d&c, number of years since previous Cesarean, gestation week and estimated fetal weight. Of the features in Figures 1 and 2, LUS full thickness feature was excluded since including both LUS full thickness and LUS myometrium thickness, which are highly correlated, degraded the performance of the ML algorithms (this issue is called multicollinearity in the literature).

**Table 3:** Performance of the ML algorithms in terms of different metrics (LUS full thickness excluded).

| Method | CRR | F-score | MCC | PRC | ROC |
| --- | --- | --- | --- | --- | --- |
| Logistic regression | 0.943 | 0.941 | 0.544 | **0.966** | 0.949 |
| Multilayer perceptron | 0.953 | 0.953 | 0.655 | 0.963 | **0.965** |
| Support vector machine | **0.956** | 0.955 | 0.659 | 0.937 | 0.816 |
| Naive Bayes | **0.956** | **0.956** | **0.672** | 0.965 | 0.950 |
| Random forest | 0.946 | 0.946 | 0.594 | 0.959 | 0.944 |

When LUS full thickness feature is replaced with LUS myometrium thickness in the model, the performance of the ML methods change as given in Table 4. It can be observed that the overall performance of the methods decrease slightly when LUS full thickness is used instead of LUS myometrium thickness.

**Table 4:** Performance of the ML algorithms in terms of different metrics (LUS myometrium thickness excluded).

| Method | CRR | F-score | MCC | PRC | ROC |
| --- | --- | --- | --- | --- | --- |
| Logistic regression | 0.950 | 0.948 | 0.610 | **0.970** | 0.965 |
| Multilayer perceptron | 0.940 | 0.938 | 0.528 | 0.969 | **0.966** |
| Support vector machine | **0.953** | 0.951 | 0.628 | 0.932 | 0.794 |
| Naive Bayes | **0.953** | **0.952** | **0.641** | **0.970** | 0.954 |
| Random forest | 0.934 | 0.932 | 0.478 | 0.963 | 0.946 |

In order to assess the importance of LUS sonography in predicting uterine dehiscence, the ML algorithms were run without either of the LUS measurements. The resulting performance of the ML methods with the features of parity, gravida, contraction, dilation, years since previous Cesarean, gestation week and estimated fetal weight was given in Table 5. It can be observed that the performance of the ML algorithms significantly degrade when LUS measurements were not taken into account.

**Table 5:** Performance of the ML algorithms in terms of different metrics (both LUS measurements excluded).

| Method | CRR | F-score | MCC | PRC | ROC |
| --- | --- | --- | --- | --- | --- |
| **Logistic regression** | 0.924 | 0.901 | 0.160 | 0.912 | 0.730 |
| **Multilayer perceptron** | 0.909 | 0.895 | 0.136 | 0.904 | 0.702 |
| **Support vector machine** | **0.927** | 0.893 | 0.001 | 0.865 | 0.500 |
| **Naive Bayes** | 0.905 | **0.912** | **0.399** | **0.924** | **0.780** |
| **Random forest** | 0.918 | 0.901 | 0.172 | 0.899 | 0.678 |

## 4. Comment

### Principal Findings

In this study, the mean value of the dehiscence group’s age was found to be higher than that of the no-dehiscence group. No significant difference was found between the groups’ BMI as well as the number of abortions, abortions after Cesarean and dilation-curettage. The number of previous Cesareans was found to be higher in the dehiscence group. Contractions in MVUs and monitorization duration was found to be not significant between the groups, while effacement percentage was found to be higher in the dehiscence group.

No significant difference was observed between the two groups in terms of smoking, dilation-curettage, gestation week, years since last Cesarean, presentation and estimated weight of the fetus. It was observed that the majority of the patients in the dehiscence group had gravida higher than 3 and parity higher than 2. Contraction in CTG and dilation prevalence was higher in the dehiscence group. Most of the patients in the dehiscence group had myometrium thicknesses of less than 1mm, while most of the patients in no-dehiscence group had more than 2mms. Estimated fetal weight did not significantly differ between the two groups.

Three rounds of ML simulations were conducted with three different sets of features. It was observed that when LUS myometrium thickness or LUS full thickness was added to the features, the prediction performance of the ML algorithms increase significantly. In terms of accuracy, Naive Bayes algorithm performed 4.8% and 5.1% better when LUS full thickness and LUS myometrium thickness was used, respectively. The increases were 4% and 4.4%, 24.2% and 27.3%, 4.6% and 4.1%, 17.4% and 17% in terms of f-score, Matthews correlation coefficient, precision recall-curve area and receiver operating characteristics area, respectively. Among the ML methods tried, the Naive Bayes yielded the best classification performance, followed closely by the support vector machine.

### Results in the Context of What is Known

In the study made by Lipschuets et al. [16] where they try to predict vaginal birth after Cesarean (VBAC) with ML, the ROC value was found to be 0.745 with the data collected from first antenatal visit, and 0.793 with the added delivery unit admission features. The ROC value in our study was found to be as high as 0.966. There are three main factors that result in this difference. First, [16] tries to predict VBAC while our study tries to predict uterine defects. Second, [16] uses parity, age, gestational week, previous delivery newborn weights, previous VBACs, dilation and presentation as features. In this work we showed that except parity, these features have little-to-no effect on the performance of the ML algorithms (see Figures 1 and 2). Lastly, we used boosting methods (gradient boosting) to increase the performance of the ML algorithms.

### Clinical Implications

This study shows that ultrasonographic measurement of LUS can be used to aid the assessment of the risk of uterine rupture. Hence, the number of untimely Cesarean operations and its complications can be reduced.

### Research Implications

Our study demonstrated that the performance of ML algorithms are sufficient to be used in clinical applications. With a bigger dataset, more complex and fine-tuned models (such as deep learning and reinforcement learning) can be used to achieve greater prediction performances.

### Strengths and Limitations

This study was conducted on a smaller dataset compared to other (nonmedical) applications of ML. Also, it suffers from imbalance (7.3% dehiscence vs. 92.7% no-dehiscence), a typical limitation of medical datasets. However, completeness of the data and high number of features (22) collected in the dataset helped to increase the performance of the ML algorithms.

## Conclusion

In conclusion, ML methods can be used to predict uterine dehiscence and hence possible rupture. Hence, the complications caused by untimely Cesarean operations can be limited. Ultrasonographic measurement of the LUS significantly increases the prediction performance of the algorithms.

## Data Availability

All data produced are available online at https://doi.org/10.17632/9hsrxcb2d2.1

